# Antibody seroprevalence and rate of asymptomatic infections with SARS-CoV-2 in Austrian hospital personnel

**DOI:** 10.1101/2021.02.01.21250898

**Authors:** Iris Leister, Elisabeth Ponocny-Seliger, Herwig Kollaritsch, Peter Dungel, Barbara Holzer, Johannes Grillari, Heinz Redl, Ivo Ponocny, Claudia Wilfing, Ludwig Aigner, Markus Exner, Michaela Stainer, Matthias Hackl, Thomas Hausner, Rainer Mittermayr, Wolfgang Schaden

**Affiliations:** Institute of Molecular Regenerative Medicine, Paracelsus Medical University, Salzburg, Austria; Spinal Cord Injury and Tissue Regeneration Center Salzburg (SCI-TReCS) and ParaMove, Paracelsus Medical University, Salzburg, Austria; Institute for Quantitative Methods, Sigmund Freud Private University, Vienna, Austria; Institute of Specific Prophylaxis and Tropical Medicine, Medical University of Vienna, Vienna, Austria; Ludwig-Boltzmann Institute for Experimental and Clinical Traumatology, Vienna, Austria; Austrian Cluster for Tissue Regeneration, Vienna, Austria; Austrian Agency for Health and Food Safety, Mödling, Austria; Institute of Molecular Biotechnology, Department of Biotechnology, BOKU – University of Natural Resources and Life Sciences, Vienna, Austria; Department for Applied Statistics and Economics, Modul University Vienna, Vienna, Austria; Labors.at GmbH, Vienna, Austria; TAmiRNA GmbH, Vienna, Austria; AUVA Trauma Center Vienna, Vienna, Austria; AUVA Headquarter, Vienna, Austria

**Keywords:** severe acute respiratory syndrome coronavirus 2, SARS-CoV-2, COVID-19 diagnostic testing

## Abstract

**Context:** On March 11, the World Health Organization (WHO) announced the current corona virus disease 2019 (COVID-19) outbreak as a pandemic. The first laboratory-confirmed case of COVID-19 in Austria was announced on February 27, 2020. Since then, the incidence of infection followed an exponential increase until a complete lockdown in March 2020. Thereafter easing of restrictions was gradually introduced and until mid-August daily infections remained mostly below 5 per 100.000 population.

**Objectives:** The aims of this study are to determine i) how many employees in Austrian trauma hospitals and rehabilitation facilities have virus specific IgG and IgM, and/or neutralizing antibodies against SARS-CoV-2, ii) how many are active virus carriers (symptomatic and asymptomatic) during the study, iii) the antibody decline in seropositive subjects over a period of around six months, and iv) the utility of rapid antibody tests for outpatient screening.

**Study Design:** Open uncontrolled observational cross-sectional study.

**Setting/Participants:** A total of 3301 employees in 11 Austrian trauma hospitals and rehabilitation facilities of the Austrian Social Insurance for Occupational Risks (AUVA) participated in the study.

**Study Interventions and Measures:** Rapid antibody tests for SARS-CoV-2 specific IgG and IgM antibodies, and RT-PCR tests based on oropharyngeal swab samples, as well as laboratory-based antibody tests using ELISA/PRNT were performed. The tests were conducted twice, with an interval of 42.4±7.7 (Min=30, Max=64) days. Additionally, participants filled out a questionnaire including questions related to personal health, traveling activities, living situation, as well as inquiries of symptoms and comorbidities. Antibody positive tested participants were re-tested with ELISA/PRNT tests at a third time point on average 188.0±12.8 days after their initial test.

**Results:** In our study cohort, only 27 out of 3301 participants (0.81%) had a positive antibody test at any time point during the study confirmed via neutralization test. Among participants who had positive test results in either of the antibody tests, 50.4% did not report any symptoms consistent with common manifestations of COVID-19 during the study period or within the preceding six weeks. In the group who tested positive during or prior to study inclusion the most common symptoms of an acute viral illness were rhinitis (21.9%), and loss of taste and olfactory sense (21.9%).

The rapid antibody test was generally more sensitive based on serum (sensitivity=86.6%) as compared to whole blood (sensitivity=65.4). Concerning both ELISA tests overall the Roche test detected 24 (sensitivity=88.9%) and the Diasorin test 22 positive participants (sensitivity=81.5%).

In participants with a positive PRNT, a significant decrease in PRNT concentration from 31.8±22.9 (Md=32.0) at T1 to 26.1±17.6 (Md=21.3) at T2 to 21.4±13.4 (Md=16.0) at T3 (χ^2^=23.848, df=2, p<0.001) was observed (χ^2^=23.848, df=2, p<0.001) – with an average time of 42.4±7.7 days between T1 and T2 and 146.9±13.8 days between T2 and T3.

**Conclusions:** During the study period (May 11^th^ – December 21^th^) only 0.81% were tested positive for antibodies in our study cohort. The antibody concentration decreases significantly over time with 14.8% (4 out of 27) losing detectable antibodies.

## Introduction

The novel severe acute respiratory syndrome coronavirus 2 (SARS-CoV-2) was first reported in December 2019 in Wuhan, China. On March 11, the World Health Organization (WHO) announced the current “corona virus disease 2019” (COVID-19) outbreak as a pandemic. The first laboratory-confirmed case of COVID-19 in Austria was announced on February 27, 2020. The incidence of infection follows an exponential growth, and the mean basic reproduction number was estimated to range from 2.24 to 3.58.^1^ After a complete lockdown in March/April, the epidemiologic situation in Austria remained stable with daily infection numbers well below 5/100.000 population until mid of August. Recently the numbers of confirmed cases continue to increase daily indicating a second wave of infections.^2^

The incubation period, defined as the time from infection to symptom onset, falls within the range of 2–14 days with 95% confidence and has a mean of around 5 days. ^3^ Backer et al. found similar results and reported an estimated mean incubation period of 6.4 days (95% CI= 5.6–7.7), ranging from 2.1 to 11.1 days (2.5th to 97.5th percentile).^4^

It has been reported that some individuals, particularly persons of younger age, infected with SARS-CoV-2 remain asymptomatic while real-time reverse transcription polymerase chain reaction (RT-PCR) tests were positive.^5,6^ Also, comparable levels of viral load in upper respiratory specimens were found in asymptomatic individuals and symptomatic patients.^7^ Asymptomatic or pre-symptomatic SARS-CoV-2 virus carriers represent a considerable pool for transmission of infection considering the communicable period is reported to be up to three weeks.^8–11^ However, the proportion of asymptomatic cases remains unclear, estimations from studies vary between 18% and 33.3%.^12^ There is evidence that this group of persons may transmit the infection for a period of at least one week although remaining PCR-positive for several weeks.^13^ This is particularly important in health care settings where patients and employees are at risk for exposure and infection.^14^

RT-PCR of the viral nucleic acid can also yield to false-negative results.^5,15–17^ False negative rates of up to 20% have been reported leading to failure to quarantine infected individuals.^18–20^ Additional serological testing of virus specific IgG and IgM antibodies is recommended because antibodies represent longer lasting markers of infection with SARS-CoV-2 in contrast to methods of pathogen detection, with the latter being detectable only transiently at the time of pathogen presence at sites where diagnostic material is collected.^5,21^

It is assumed, that most individuals who are infected with SARS-CoV-2 develop antibodies within three weeks after onset of Covid-19 symptoms.^14,22,23^ However, some studies reported a rapid waning of antibody titers indicating that humoral immunity against SARS-CoV-2 may be very short-lived, particularly in patients with asymptomatic infection.^21,24^ Among routine antibody tests ELISA tests for antibodies directed against RBD (receptor binding domain) of SARS-CoV spike protein are considered to be a kind of "gold standard” to detect protective antibodies whilst IgA and IgG ELISA tests against viral nucleoproteins have only diagnostic value.^25^ In addition, tests for detection of neutralizing antibodies are considered to be reference for all kinds of routine tests.

Sensitivity rates as well as specificity rates of recently evolved and currently available rapid antibody tests are not yet reliable enough.^16,26,27^ Additionally, many of those tests are not entirely specific for the SARS-CoV-2 virus, because of a cross-reactivity to other human SARS viruses like HCoV-OC43, HCoV-HKU1, HCoV-229E and HCoV-NL63.^23^ Therefore, it is recommended to evaluate the antibody status via laboratory based enzyme linked immunosorbent assay (ELISA) and / or plaque reduction neutralization test (PRNT).^25^

The aims of this study were to determine i) how many employees in Austrian trauma hospitals and rehabilitation facilities have virus specific IgG and IgM, and/or neutralizing antibodies against SARS-CoV-2, indicating that they have recovered from infection with or without having shown symptoms at any time, ii) how many were actively infected during the study period and did not show COVID-19 associated symptoms while included in the study, iii) the antibody decline in seropositive subjects over a period of around six months, and iv) the utility of rapid antibody tests for outpatient screening.

## Methods

This is an open, uncontrolled, three-time cross-sectional study.

### Study population

All employees in Austrian trauma hospitals and rehabilitation facilities of the Austrian Social Insurance for Occupational Risks (AUVA) have been invited to participate in the study. In total, 3301 hospital employees were enrolled in eleven participating hospitals and rehabilitation facilities in Austria. The mean(SD) age in the cohort was 43.8 (11.9) years, 2246 (68%) are female, 1049 (31.8%) are male, and 6 (0.2%) refused to disclose their gender. Written informed consent has been obtained from all study participants.

### Study procedures

The study procedures are depicted in figure 1. A rapid antibody test (chip based) for SARS-CoV-2 specific IgG and IgM antibodies, and an RT-PCR test based on oropharyngeal swab samples, as well as the ELISA / PRNT tests (if indicated) were offered to all employees in Austrian trauma hospitals and rehabilitation facilities. The rapid antibody tests and PCR tests were conducted twice, with 42.4±7.7 (Min=30, Max=64) days in between testing. The lab-based ELISA and/or PRNT was conducted only if the rapid antibody test showed a positive or questionable result or if a prior positive PCR was known. Participants who had positive antibody tests in ELISA and/or PRNT, were invited for a third blood sample collection 186.8±11.5 days after their initial positive result to determine antibody kinetics. Individuals who agreed to participate in the study were asked to fill out an additional questionnaire to identify special risk factors which consisted of the following items:

**Figure 1:**
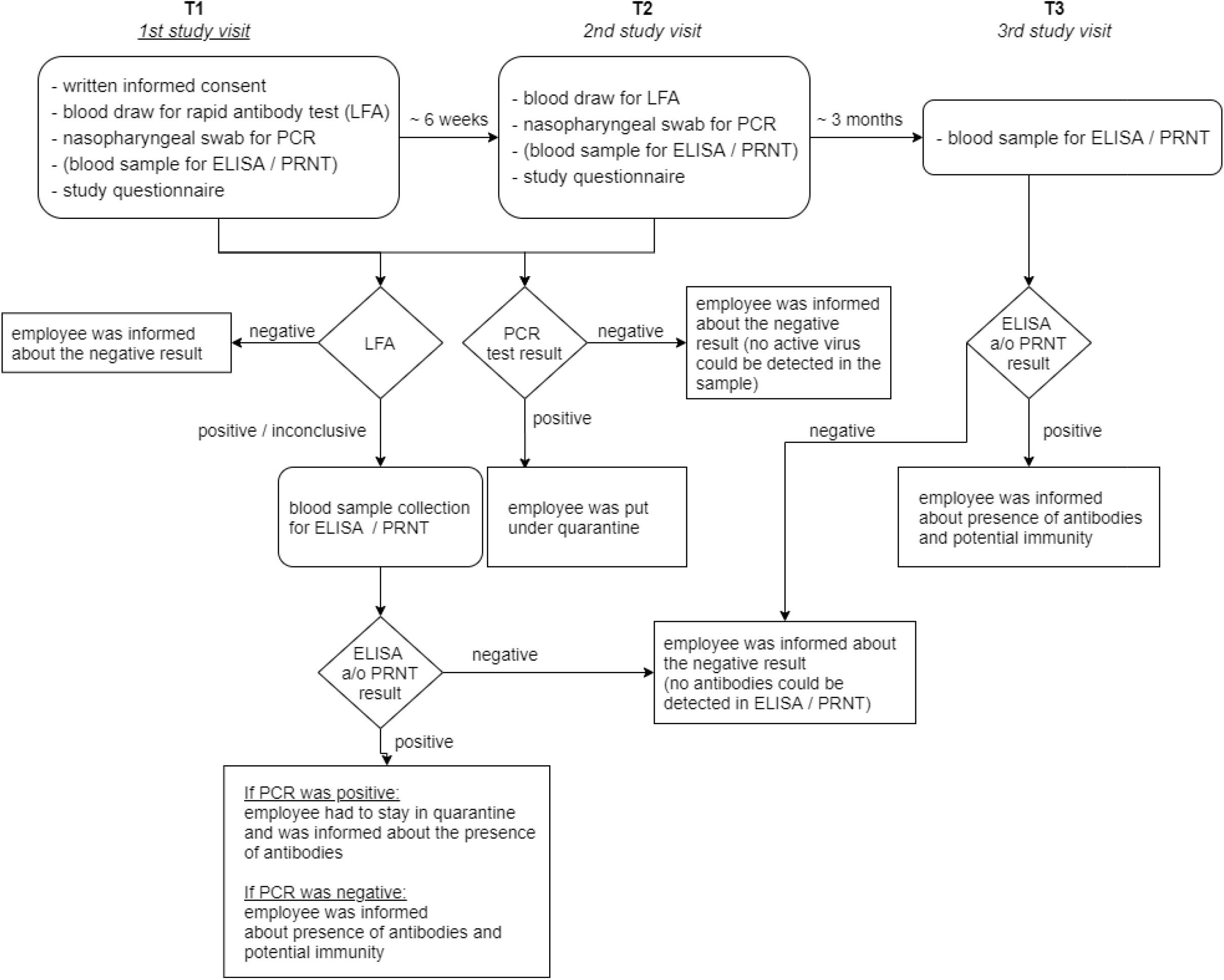
Study procedures. Figure created with draw.io

- demographics (name, contact details, gender, age, height, bodyweight)
- code of the hospital or rehabilitation facility
- if the participant is a smoker including a quantification of smoking habit
- if the person had been tested for SARS-CoV-2 previously (with either an antibody test or a RT-PCR) including the test result
- survey flu like symptoms (at present or within the previous six weeks)
  - fever
  - cough
  - dyspnea
  - nasal congestion
  - sore throat
  - headaches
  - myalgia
  - impaired sense of smell or taste
  - diarrhea
- questions related to the living situation
  - how many people live in the same household
  - how many adolescents below the age of 15 years live in the same household
  - inquiry of flu like symptoms within the family members / housemates
- questions related to commuting
  - use of public transportation
  - by car
  - by bike
  - walking
- survey on traveling activities within the previous six weeks
  - eastern Austria
  - western Austria
  - Italy
  - within Europe
  - outside of Europe
- attendance on large-scale events with more than 100 people within the last six weeks
- job description (divided into 25 categories: medical doctors, nurses, administrative staff, cleaning staff, medical-technical staff, laboratory staff, etc.)
- working hours within the last four weeks and periods of absence
- inquiry of comorbidities
  - hypertension
  - diabetes mellitus
  - coronary artery disease
  - chronic obstructive pulmonary disease (COPD)
  - chronic kidney disease
- inquiry of immunosuppression and allergies
- test results of the IgG and IgM antibody test
- test result of the RT-PCR test based on oropharyngeal swab samples
- test result of the ELISA / PRNT

The period of sample collection was from May 11th until August 23^rd^ 2020 for the PCR and LFA, and until December 4^th^ 2020 for the additional blood sample collection in subjects with positive results prior.

### Real-time reverse transcription polymerase chain reaction (RT-PCR) tests

#### RNA Isolation

Total RNA was isolated from 200µL of virus media using the commercially available Maxwell RSC Viral Total Nucleic Acid Purification Kit (Promega, AS1330) in combination with the Maxwell RSC48 system. Samples were mixed with 200µL lysis buffer, 20µL Proteinase K, and 0.5µL internal positive control (IPC, Ingenetix Viroreal SARS & SARS-CoV-2, DHUV02313×5) followed by vigorous mixing for a minimum of 10 seconds and 10 minutes incubation at room temperature. Afterwards samples were applied to Promega Maxwell cartridges and subject to automated processing on the Maxwell RSC 48 system according to the manufacturer’s instructions. Samples were purified with paramagnetic particles that serve as mobile solid phase. Total RNA was eluted in 50 µl nuclease-free water.

#### SARS-CoV-2 Screening Assay

The CE-IVD ViroReal SARS-CoV-2 & SARS Kit (Ingenetix, Austria, Cat: DHUV02313×5) was used for detection of SARS-CoV-2 nucleocapsid RNA in the clinical sample material. Probes were designed to detect the nucleocapsid protein gene (N gene) of SARS-CoV-2 as well as SARS-CoV and SARS-related coronavirus. Other beta coronaviruses are not detected with this kit. The primer and probe design chosen by this assay is not identical to the WHO design, as it intends to cover possible future changes in the virus sequence, therefore a highly conserved region in all SARS coronavirus clusters of the N gene was chosen as target region. This allows universal detection of all so far known SARS-CoV strains including SARS-CoV-2 and SARS-like CoV without discriminating between strains.

The test uses a one-step reverse transcription real-time PCR (RT-PCR) reaction. A probe-specific amplification curve at 530 nm (FAM channel) indicates the amplification of virus-specific RNA. An internal RNA positive control (RNA IPC) is detected in Cy5 channel and was used as internal PCR control for both RNA extraction as well as RT-PCR efficiency. The target for the RNA IPC is extracted together with the sample. To set up the reaction 5µL isolated RNA were mixed with 2 µL nuclease free water (NFW), 2.5µL reaction mix and 0.5µL SARS-CoV-2 & SARS Assay Mix + RNA IPC Assay Mix. RT-qPCR was performed under following conditions: RT: 50°C for 15min, Activation: 95°C for 20sec, PCR 45 cycles: 95°C for 5 sec and 60°C for 30 sec. Cycle of quantification (Cq) was calculated with the second derivative maximum method.

#### SARS-CoV-2 Confirmation Assay

Positive results from the screening assay SARS-CoV-2 were confirmed on the basis of an independent detection of the SARS-CoV-2 specific E-Gene. This protocol is based on a publication from Corman et al. “Detection of 2019 novel coronavirus (2019nCoV) by real time-RT-PCR” ^28^ using Superscript III RT-qPCR System (Thermo Fisher, Cat: 12574026). Per reaction, 3.44µL RNA were mixed with 5µL reaction mix, 0.4µL RT/Polymerase, 0.4µL 1µg/µL BSA, 0.16µL MgSO4 and 0.2µL of each E-Gene forward and reverse primer and probe. RT-qPCR is performed under following conditions: RT: 55°C for 10min, Activation: 95°C for 3min, PCR 45 cycles: 95°C for 15 sec and 58°C for 30 sec. Cq-value was calculated with the second derivative maximum method. Sequences of the used primers and probes are shown below.

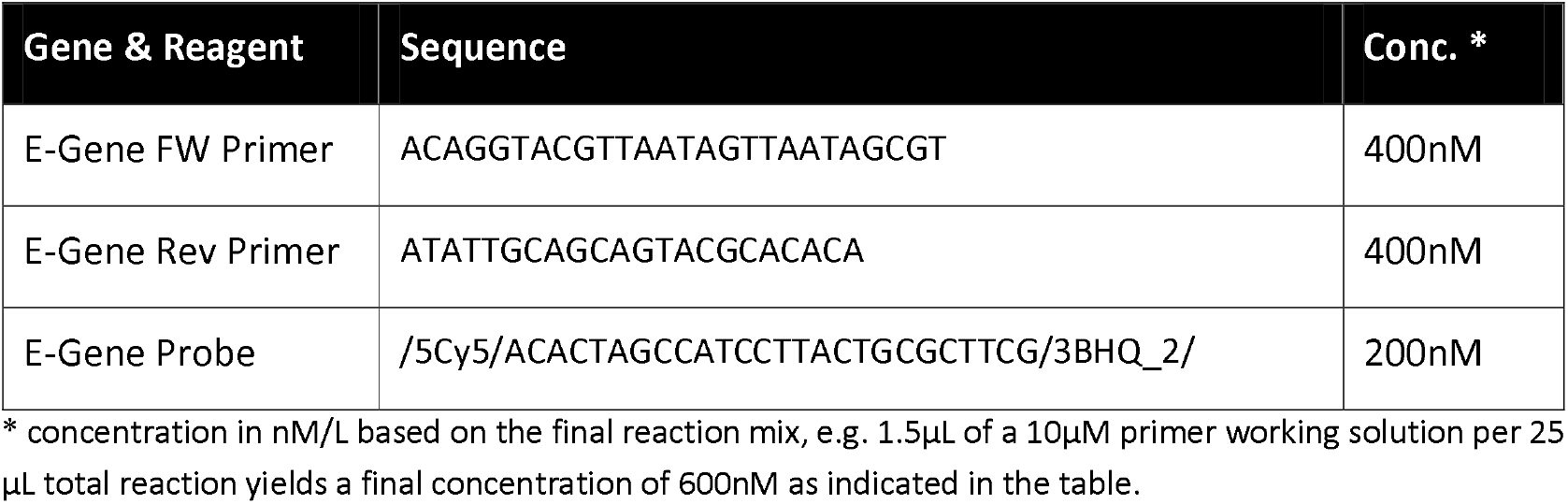

### Rapid antibody test: SARS-CoV-2 Antibody lateral flow assay (LFA)

LFAs were performed according to the instructions-for-use (TAmiRNA GmbH, Austria). In brief, all test components were brought to room temperature. Sera or plasma aliquots were completely thawed before testing, or whole blood was freshly collected using safety lancets. The test cassette was removed from the sealed pouch and the required amount of sample (20 µl serum/plasma or 20 µl whole blood) was pipetted into the specimen well using clean disposable pipets. Immediately after the sample was applied to the sample inlet, three drops of sample buffer were applied to the buffer well. Results were read within the specified time window of 15 minutes.

### Laboratory-based ELISA (Roche and Diasorin)

SARS-CoV-2 specific antibodies were measured from serum samples with two assays: (i) a qualitative eCLIA assay targeting pan-immunoglobulin (IgM, IgA, IgG) antibodies against the nucleoprotein (N-Ag) (Roche Elecsys Anti-SARS-CoV-2) on a cobas e601 analyzer. Samples with a cutoff index COI <1.0 are classified as non-reactive. (ii) a quantitative CLIA assay targeting IgG antibodies against the S1/S2 protein (Diasorin LIAISON SARS-CoV-2 S1/S2 IgG) on a LIAISON XL analyzer. Results are expressed in arbitrary units (AU/ml). Samples with <12,0 AU/ml are classified as negative; 12,0 ≤ x < 15,0 as borderline and ≥ 15,0 as positive.

According to the manufacturer, for the Roche Elecsys Anti-SARS-CoV-2 assay sensitivity rates range between 65.5% and 100% depending on the time between diagnosis and sample collection (65.5% 0-6 days, 88.1% 7-13 days, and 100% more than 14 days after diagnosis, respectively).

Sensitivity rates for the Diasorin LIAISON SARS-CoV-2 S1/S2 IgG range between 25% and 97.4% and are also dependent on the time since diagnosis via RT-PCR according to the manufacturer (25% less than 5 days, 90.4 % 5-15 days, and 97.4% more than 15 days after diagnosis).

### Neutralization test

#### Cell lines and viruses

Vero 76 clone E6 cells (CCLV-RIE929, Friedrich-Loeffler-Institute, Riems, Germany) were cultured in minimum essential medium Eagle (E-MEM) with Hank’s balanced salt solution (HBSS) (BioWhittaker, Lonza, Szabo Scandic, Austria), supplemented with 10% fetal bovine serum (Corning, Szabo Scandic, Austria) (FBS) and were used to titrate virus preparations and for the neutralization assays. Vero E6 TMPRSS-2 (provided by Stefan Pöhlmann; Deutsches Primatenzentrum, Göttingen, Germany) - initially described in Hoffmann et al. - were cultured in Dulbecco’s modified Eagle’s medium (DMEM) with 10% fetal bovine serum (FBS) and used for virus passaging and isolating infectious virus from clinical samples.^29^ The virus used for the neutralization assay was originally isolated from a clinical specimen (NP swab), taken in mid-March 2020 from a 25-year old male patient in Lower Austria and further passaged twice on Vero E6 TMPRSS-2 cells.

#### Neutralization assay

The neutralization assay was set up in flat-bottom 96-well tissue culture plates. Human sera were heat-treated for 30 min at 56°C and diluted 1 to 4 in triplicates in serum-free DMEM medium as starting point for the assay. Two-fold serially diluted sera were incubated with an equal volume of 50 μl SARS-CoV-2 at a minimum of 2,000 tissue culture infectious dose 50% (TCID50)/ml) for 90 min at 37°C. Next, 25,000 Vero 76 clone E6 cells were added to the serum/virus mixture in each well in a volume of 100 µl in EMEM, supplemented with 10% FBS and incubated for 4 days at 37°C, 5% CO2 in a humidified incubator. The CPE in every well was scored under an inverted optical microscope and the reciprocal of the highest serum dilution that protected more than 50% of cells from CPE was taken as the neutralizing titer.

PRNT was used as a reference in this study because neutralization assays are proposed to have the highest validity for CoV serology.^23^

### Statistical analysis

Statistical analysis was performed using SPSS statistical software (version 26, SPSS Inc., Chicago, Illinois, USA). Figures were compiled with Biorender.com, draw.io, and Microsoft Excel (Microsoft Corp., Redmond, WA, USA). Results of the PCR and rapid antibody test (IgG and IgM) were included in the eCRF at both time points together with all questionnaire data. If an ELISA and / or PRNT was indicated, results of these tests have also been included in the eCRF.

The sensitivity of the rapid antibody and ELISA test was calculated with respect to the neutralization test for all three time points separately. As blood was only sampled in case of a positive prior PCR or a positive or questionable rapid antigen test, sensitivity estimations are biased in favor of these tests and specificity cannot be determined.

Data are analyzed exploratively; for comparison of groups concerning sociodemographic and medical variables chi-square-tests and t-tests depending on the scale are applied. Reduction in antibody concentration between time points is tested via Friedman test and Spearman correlation is used to evaluate dependencies between reduction in antibody concentration, age of the participants, and days between T1/T2 and T2/T3, respectively.

## Results

Details on demography and occupation of the study participants are summarized in table 1. At first time point (T1), a total of n_1_=3301 hospital employees could be enrolled whereby sociodemographic variables (see Table 1) were comparable to the total AUVA population. At the second test time point (T2), which was on average 40.1±9.8 days later, a reduced number of n_2_=2941 could be re-tested by means of a PCR test.

**Table 1.**
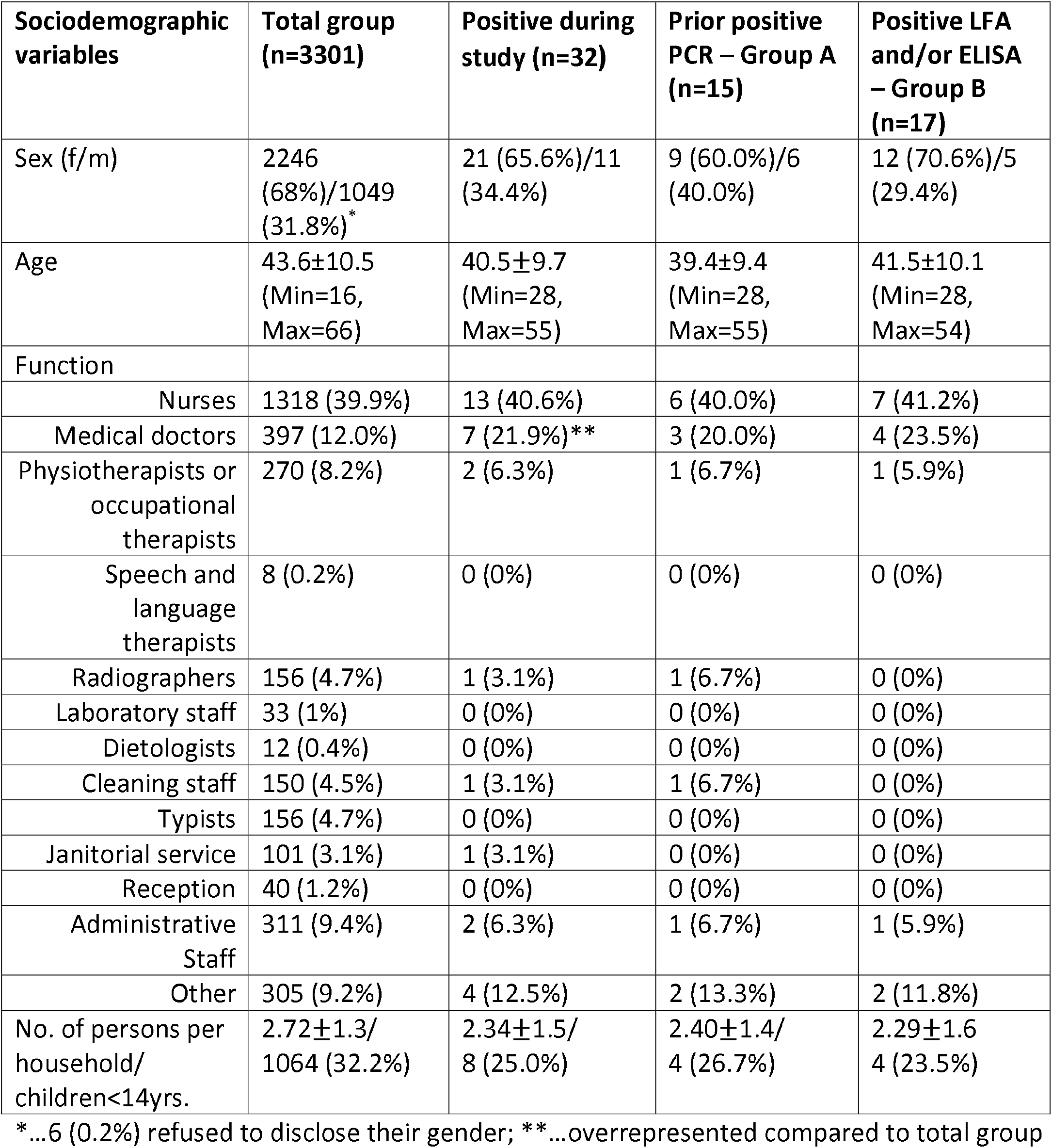
Sociodemographic Variables – All Groups

| **Sociodemographic variables** | **Total group (n=3301)** | **Positive during study (n=32)** | **Prior positive PCR – Group A (n=15)** | **Positive LFA and/or ELISA – Group B (n=17)** |
| --- | --- | --- | --- | --- |
| Sex (f/m) | 2246 (68%)/1049 (31.8%)* | 21 (65.6%)/11 (34.4%) | 9 (60.0%)/6 (40.0%) | 12 (70.6%)/5 (29.4%) |
| Age | 43.6±10.5 (Min=16, Max=66) | 40.5±9.7 (Min=28, Max=55) | 39.4±9.4 (Min=28, Max=55) | 41.5±10.1 (Min=28, Max=54) |
| Function |  |  |  |  |
| Nurses | 1318 (39.9%) | 13 (40.6%) | 6 (40.0%) | 7 (41.2%) |
| Medical doctors | 397 (12.0%) | 7 (21.9%)** | 3 (20.0%) | 4 (23.5%) |
| Physiotherapists or occupational therapists | 270 (8.2%) | 2 (6.3%) | 1 (6.7%) | 1 (5.9%) |
| Speech and language therapists | 8 (0.2%) | 0 (0%) | 0 (0%) | 0 (0%) |
| Radiographers | 156 (4.7%) | 1 (3.1%) | 1 (6.7%) | 0 (0%) |
| Laboratory staff | 33 (1%) | 0 (0%) | 0 (0%) | 0 (0%) |
| Dietologists | 12 (0.4%) | 0 (0%) | 0 (0%) | 0 (0%) |
| Cleaning staff | 150 (4.5%) | 1 (3.1%) | 1 (6.7%) | 0 (0%) |
| Typists | 156 (4.7%) | 0 (0%) | 0 (0%) | 0 (0%) |
| Janitorial service | 101 (3.1%) | 1 (3.1%) | 0 (0%) | 0 (0%) |
| Reception | 40 (1.2%) | 0 (0%) | 0 (0%) | 0 (0%) |
| Administrative Staff | 311 (9.4%) | 2 (6.3%) | 1 (6.7%) | 1 (5.9%) |
| Other | 305 (9.2%) | 4 (12.5%) | 2 (13.3%) | 2 (11.8%) |
| No. of persons per household/ children<14yrs. | 2.72±1.3/ 1064 (32.2%) | 2.34±1.5/ 8 (25.0%) | 2.40±1.4/ 4 (26.7%) | 2.29±1.6 4 (23.5%) |

\*...6 (0.2%) refused to disclose their gender; \*\*...overrepresented compared to total group

Within the study cohort, 334 persons (10.1%) had comorbidities that are considered to be associated with a more severe course of disease in Covid-19 (see table 2). Among the participants with comorbidities, 257 (7.8%) had hypertension, 41 (1.2%) had diabetes mellitus, 19 (0.6%) had coronary artery disease, 43 (1.3) had COPD, and 14 (0.4%) had chronic kidney disease. These 334 individuals, together with an additional 31 (0.9%) immunocompromised persons are considered a risk group in the event of infection with SARS-CoV-2. Furthermore 1116 (33.8%) participants reported to have allergies. (table 2) A total of 769 (23.3%) employees are smokers, whereby 513 (15.5%) smoke between six and 15 cigarettes per day, 212 (6.4%) smoke more than 15 cigarettes per day, and 44 (1.3%) stated to use other products containing nicotine. Chi-squared analysis revealed that in individuals who smoke, the incidence of flu-like symptoms within six weeks before study enrollment was significantly higher (11%) compared to non-smokers (9%, p < 0.001, Cramer V = 0.71).

**Table 2.** Health Variables – All Groups

| Health Variables | Total group<br>(n=3301) | Positive during<br>study (n=32) | Prior positive<br>PCR – Group A<br>(n=15) | Positive LFA<br>and/or ELISA<br>– Group B<br>(n=17) |
| --- | --- | --- | --- | --- |
| Hypertension | 257 (7.8%) | 3 (9.4%) | 1 (6.7%) | 2 (11.8%) |
| Diabetes Mellitus | 41 (1.2%) | 0 (0%) | 0 (0%) | 0 (0%) |
| Coronary Artery<br>Disease | 19 (0.6%) | 1 (3.1%) | 1 (6.7%) | 0 (0%) |
| COPD | 43 (1.3%) | 0 (0%) | 0 (0%) | 0 (0%) |
| Chronic Kidney<br>Disease | 14 (0.4%) | 0 (0%) | 0 (0%) | 0 (0%) |
| Immunocompromised | 31 (0.9%) | 1 (3.1%) | 0 (0%) | 1 (5.9%) |
| Allergies | 1116 (33.8%) | 14 (43.8%) | 8 (53.3%) | 6 (35.3%) |
| Smoker | 769 (23.3%) | 5 (15.6%) | 3 (20.0%) | 2 (11.8%) |

### Active virus carriers during the study period

Only one person had a positive PCR test result at T1 on May 12^th^. This participant was a medical doctor, who already tested positive 60 days before study inclusion and is therefore allocated to group A in our analysis.

### Participants with positive test results

A total of n_P_=32 participants were tested SARS-CoV-2 positive at any time point either prior to the study (i.e. prior positive PCR-Test = Group A) or were tested positively for antibodies during the study (positive or questionable/inconclusive rapid antibody test and/or positive ELISA (Roche and/or Diasorin) = Group B). (Sociodemographic and health characteristics of these groups can be found in Table 1 and Table 2, respectively.)

Nineteen (59.4%) out of 32 test-positive participants reported no symptoms of an acute viral illness within six weeks prior to study enrollment as well as during the study period. PRNT was performed if any antibody test was positive. Due to limited serum sample availability in 4 out of 32 participants, PRNT was performed on 28 sera with 25 out of which neutralizing activity was observed (one test was questionable). One participant lost neutralizing antibodies between T1 and T2 and another three participants lost them between T2 and T3.

Concerning clinical signs, rhinitis (21.9%) and loss of taste and olfactory sense were the most prominent symptoms (21.9%; table 3) in the group who tested positive at any time during or prior to study inclusion.

**Table 3:**
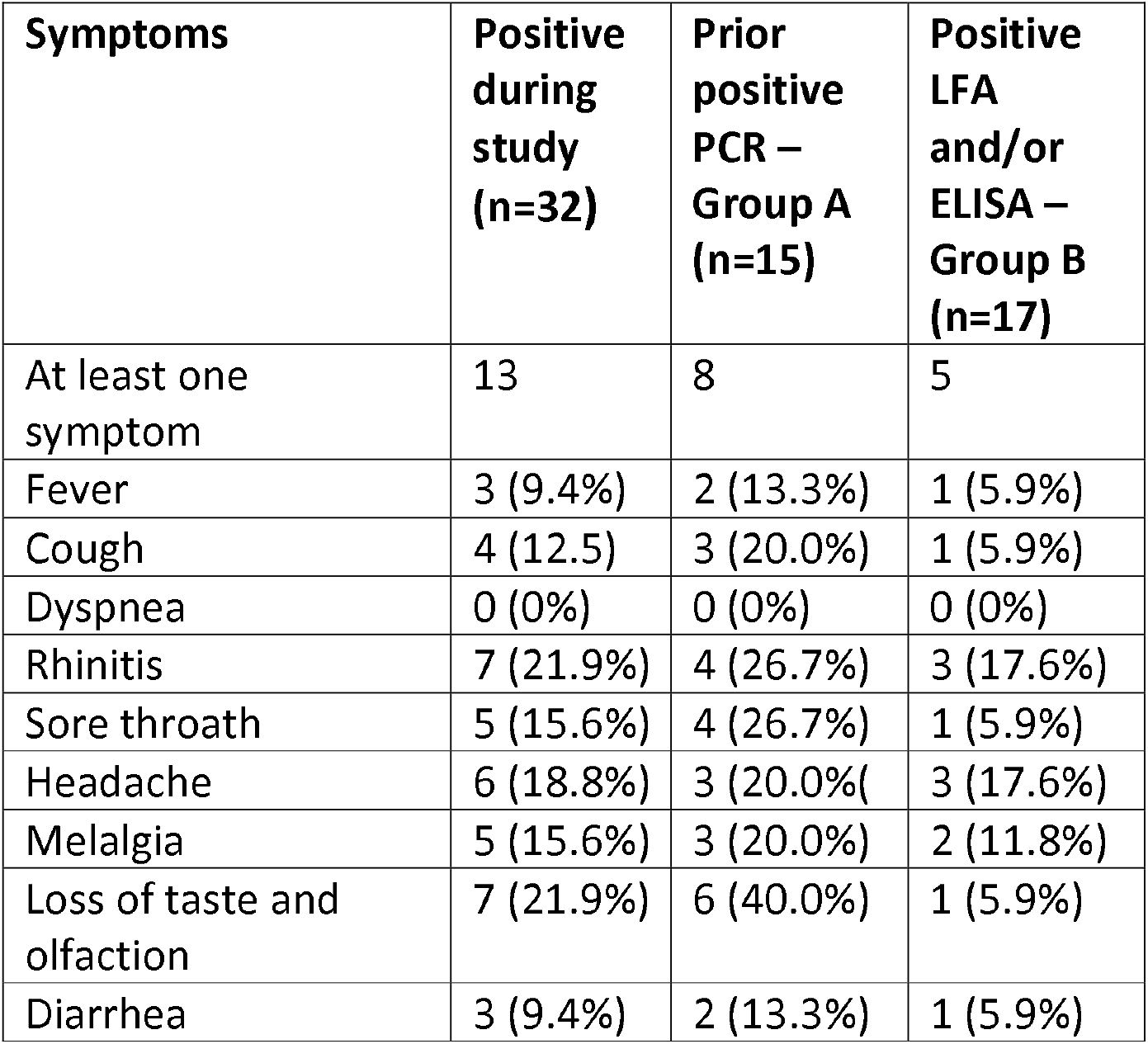
Symptoms – All Groups

| Symptoms | Positive<br>during<br>study<br>(n=32) | Prior<br>positive<br>PCR –<br>Group A<br>(n=15) | Positive<br>LFA<br>and/or<br>ELISA –<br>Group B<br>(n=17) |
| --- | --- | --- | --- |
| At least one<br>symptom | 13 | 8 | 5 |
| Fever | 3 (9.4%) | 2 (13.3%) | 1 (5.9%) |
| Cough | 4 (12.5) | 3 (20.0%) | 1 (5.9%) |
| Dyspnea | 0 (0%) | 0 (0%) | 0 (0%) |
| Rhinitis | 7 (21.9%) | 4 (26.7%) | 3 (17.6%) |
| Sore throat | 5 (15.6%) | 4 (26.7%) | 1 (5.9%) |
| Headache | 6 (18.8%) | 3 (20.0%) | 3 (17.6%) |
| Melalgia | 5 (15.6%) | 3 (20.0%) | 2 (11.8%) |
| Loss of taste and<br>olfaction | 7 (21.9%) | 6 (40.0%) | 1 (5.9%) |
| Diarrhea | 3 (9.4%) | 2 (13.3%) | 1 (5.9%) |

### Group A: Participants with a positive PCR test prior to study inclusion

Fifteen employees had positive PCR test results for acute SARS-CoV-2 infection prior to study inclusion, with an average of 44.5±20.0 days between a prior positive PCR and T1 in the present study. Out of these, eight persons suffered from symptoms in the preceding six weeks.

Thirteen of these employees showed a positive neutralization test, for one participant the neutralization test result was below the threshold of a 1:4 dilution, and for another participant no blood sample could be obtained. Based on the blood samples only for seven of these participants the rapid antibody test was positive – including the person with the neutralization value below the threshold – so the sensitivity for the rapid antibody test in group A at T1 was only 46.2% (figure 3). Using the rapid antibody test with serum nine positive results (sensitivity= 69.2%) and the one participant with the neutralization value below the threshold could be detected correctly.

**Figure 2:**
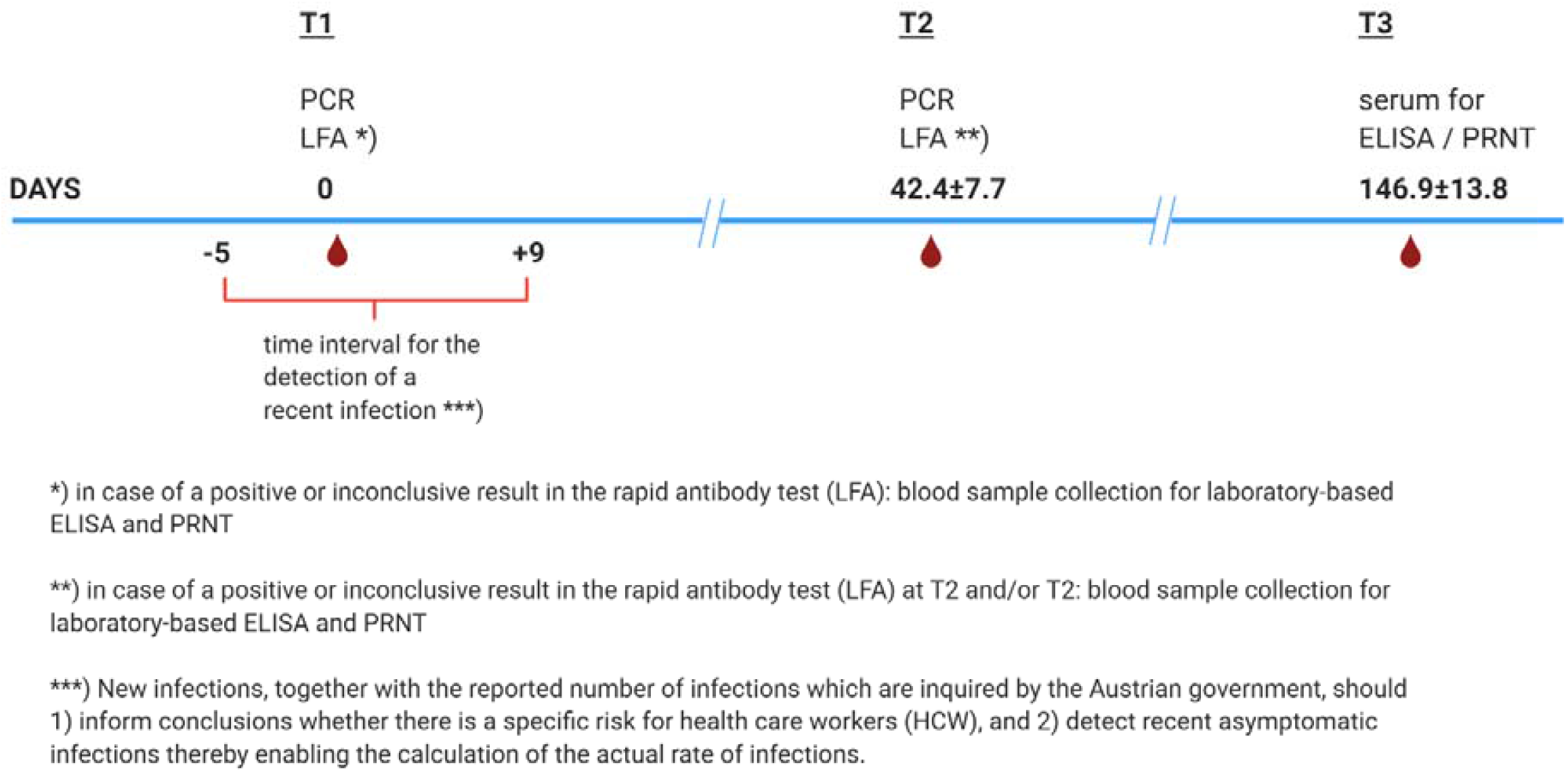
time intervals of testing. Figure created with Biorender.com

**Figure 3:**
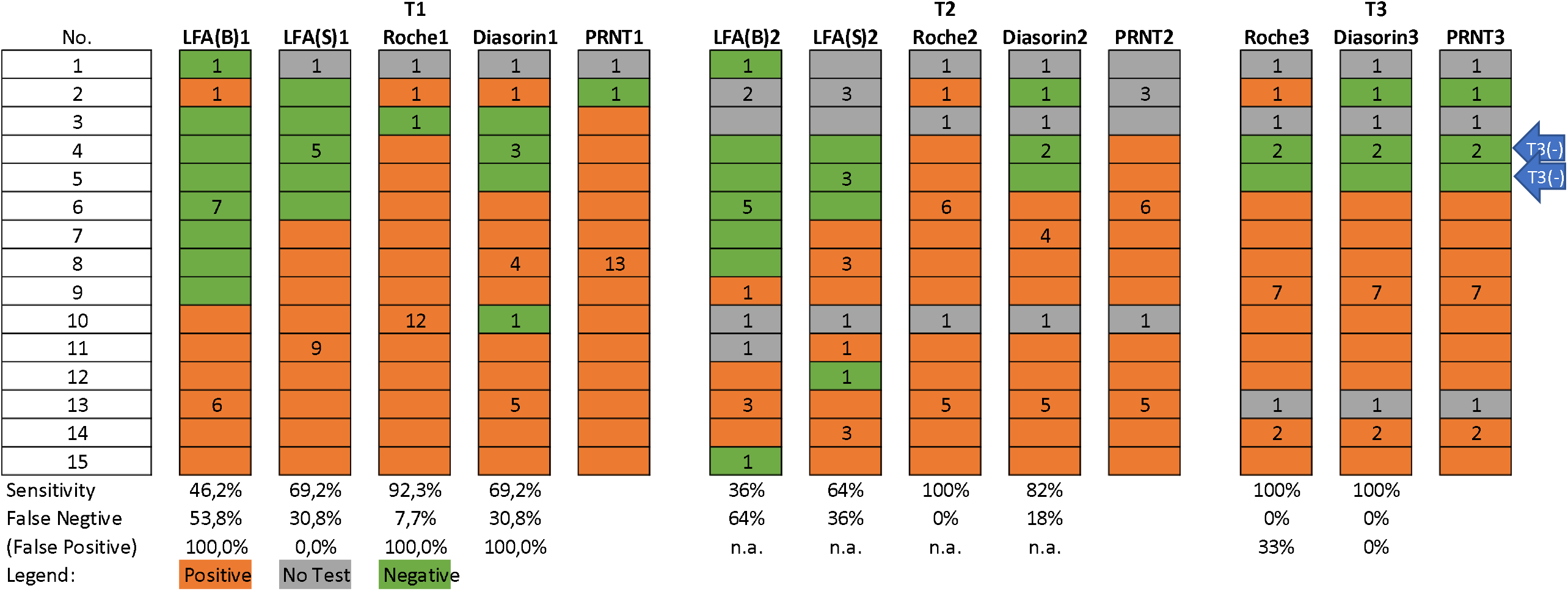
Group A – PRNT, ELISA (Roche and Diasorin) and rapid antibody test (LFA) with either whole blood (LFA(B)) or blood serum (LFA(S)) at T1, T2 and T3

A total of thirteen participants had a positive ELISA-test from Roche again including the one person with the neutralization test below the threshold, resulting in a sensitivity of 92.3% when compared with the neutralization test. Based on the ELISA-test from Diasorin ten participants were positive, again including the NT negative participant (sensitivity=69.2%). At the second timepoint (T2), a neutralization test result could only be obtained for eleven participants (all of them have tested positive at T1). The rapid antibody test based on whole blood was only positive for four of them (sensitivity = 36.0%), while based on blood serum it was positive for seven (sensitivity=64%). Concerning both ELISA tests the Roche test detected all eleven positive participants (sensitivity=100%) and the Diasorin test only nine positive participants (sensitivity=82%).

For twelve participants a neutralization test result was obtained at (T3), showing that two persons lost their antibodies between T2 and T3 (marked with T3(-) in figure 3); both ELISA tests detected all nine positive participants (sensitivity=100%), the Roche test resulted in one false positive whereas the Diasorin test detected all three negative participants correctly.

Concerning those participants where a positive neutralization test was obtained, a significant decrease of the value from 26.5±14.7 (Md=32.0) at T1 to 21.8±16.2 (Md=16.0) at T2 and to 17.8 ±9.8 (Md=16.0) at T3 could be seen (χ2=10.4, df=2, p=0.006) – with an average time between T1 and T2 of 40.3±6.1 days and between T2 and T3 of 151.6 ±8.1 days.

### Group B: Participants with a positive antibody test during the study, either rapid antibody and/or ELISA at T1 and/or T2

Seventeen employees were tested positive for antibodies during the study, either with the rapid antibody test and/or one of the ELISA tests, whereby the latter were only performed in individuals with a prior positive or questionable rapid antibody test. For 14 of these participants a neutralization test for validation could be performed. The majority was asymptomatic as only five participants reported symptoms in the preceding six weeks.

At T1, 14 persons of group B had a positive rapid antibody test based on whole blood, which could be confirmed for ten persons via neutralization test (sensitivity = 83.3%; figure 4). Using blood serum for the rapid antibody test, which was available only for 13 persons, all of them were positive – compared with the neutralization test but also one negative participant and one participant with a questionable result (sensitivity=100%).

**Figure 4:**
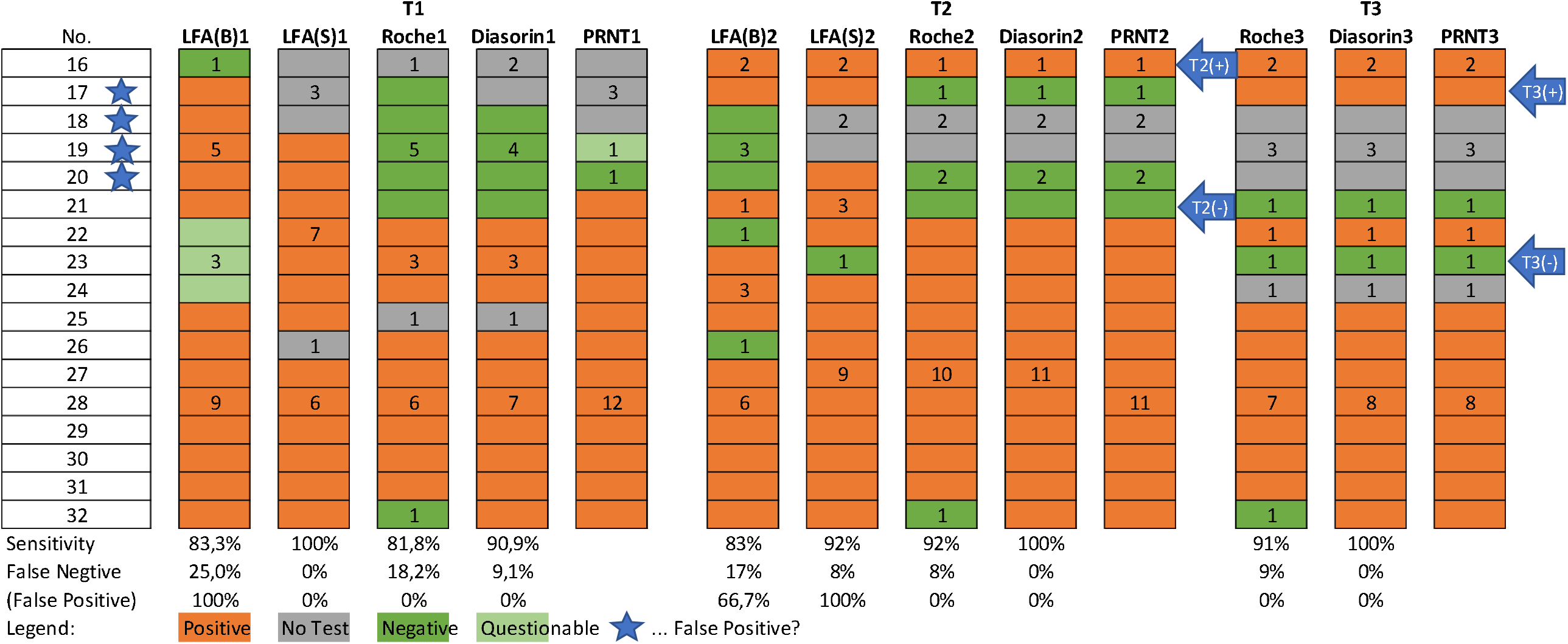
Group B – PRNT, ELISA (Roche and Diasorin) and rapid antibody test (LFA) with either whole blood (LFA(B)) or serum (LFA(S)) at T1, T2 and T3

The ELISA-Roche test was positive for nine persons (sensitivity=81.8%) and the ELISA-Diasorin test was positive for ten persons (sensitivity=90.9%) again compared to the neutralization test (figure 4). At T2 neutralization tests could be performed for fifteen persons; one additional person who had a negative PCR and negative rapid antibody test at T1 but reported on flu-like symptoms prior to T1 was tested positive at T2 (No 16, marked T2(+) in figure 4) and one person seemingly lost his/her antibodies in the 41 days after T1 (No 21, marked T2(-) in figure 4). Based on whole blood, 12 persons were tested positive with the rapid antibody test, whereby ten cases (sensitivity=83.0%) which also showed neutralization activity. Although, based on blood serum, 14 persons had positive results in the LFA, 11 of which also showed neutralization activity (sensitivity= 92.0%). The ELISA-Roche showed a sensitivity of 92% and the ELISA-Diasorin one of 100% for T2 using PRNT as reference.

Thirteen participants could be re-tested at T3 with the neutralization test showing a positive result for eleven persons. One additional participant became positive (No 17, marked with T3(+) in figure 4) which had so far only been positive in the rapid antibody test at T1 and T2, but reported flu-like symptoms between T2 and T3. Another person lost detectable antibodies (No 23, marked with T3(-) in figure 4). The ELISA-Diasorin test correctly identified all positive (sensitivity=100%) and negative participants while the ELISA-Roche test resulted in one false negative result (sensitivity=91%).

Concerning those four participants (no 17, 18, 19, 20) who are marked with an asterisk in figure 4, only the rapid antibody test was positive either at T1 and/or T2 – so it may be hypothesized that they have always been false positive in the rapid antibody test and patient no 17, who showed antibodies at T3 most probably had corona between T2 and T3. Also, in group B a significant decrease in the neutralization test value from 37.5±29.0 (Md=32.0) at T1 to 30.1±18.6 (Md=26.7) at T2 to 24.4± 15.6 (Md=28) at T3 could be seen (χ^2^=14.389, df=2,, p=0.001) – with an average time of 43.7±8.5 days, and 143.6±16.2 days between T1 / T2, and T2 / T3, respectively.

The rapid antibody test shows a low sensitivity of only 43.2% (T1)/36% (T2) based on blood and 69.2% (T1)/64% (T2) based on serum in a group of participants who had been PCR positive recently. But, if a positive or questionable rapid antibody test result was obtained, the result could be confirmed via PRNT at the same time of sample collection for 83.3% (T1)/83% (T2) based on blood and 100% (T1)/92% (T2) based on serum. Summarizing these findings, the ELISA-Roche shows higher sensitivity than ELISA-Diasorin in participants who have been positive a priori, but in validating the rapid antibody test results ELISA-Diasorin outperformed ELISA-Roche.

### Decline of antibody concentration

PRNT values showed a significant decrease from 31.8±22.9 (Md=32.0) at T1 to 26.1±17.6 (Md=21.3) at T2 to 21.4 ±13.4 (Md=16.0) at T3 (χ^2^=23.848, df=2, p<0.001) – with an average time of 42.4±7.7 days between T1 and T2 and 146.9±13.8 days between T2 and T3. Only two participants maintained their antibody titers at all three time points. Generally, the decrease in concentration is independent of the time period between T1/T2 and T2/T3, respectively (ρ_T1/T2_=.065, p_T1/T2_=0.775; ρ_T2/T3_=.042, p_T1/T2_=0.861).

### General antibody seroprevalence

No difference in terms of antibody seroprevalence was observed between male and female participants, smokers and non-smokers, the geographical area of the participating center, or the presence of comorbidities or allergies (p>0.05). Only medical doctors were slightly overrepresented (p=0.097) in the group of positive cases, and individuals who were tested positive showed somewhat more traveling activities, especially outside Europe in February/March (p=0.010). The number of persons per household was even slightly smaller (2.34±1.5 vs. 2.73±1.32; z=1.783, p=.075) in the group of persons with antibody seroprevalence with no differences according to the number of children below the age of 15 years (p>0.05). Only seven persons reported of cohabitants with flu-like symptoms at least six weeks prior to the study whereby five had symptoms themselves (the neutralization test confirmed antibody seroprevalence for six persons and for one person the results were questionable). Also, asymptomatic persons do not differ from symptomatic ones in the neutralization test values (p>.0.05). Finally, persons with positive seroprevalence do not differ according to how they reach their work place (public transport, car, bicycle or walk; p>0.05).

## Discussion

In our study cohort, only 32 out of 3301 participants (0.96%) had a positive antibody test at any time point during the study whereby results could be confirmed for 27 via neutralization test. This is remarkably lower compared to a large multicenter study in the USA, where 6% had detectable SARS-CoV-2 antibodies.^14^ Also, a large study on 61 075 residents of Spain reported a seroprevalence between 4.6 and 5% (by point-of-care test and immunoassay, respectively) where sample collection took place between April 27 and May 11, 2020.^30^ Another study by Korth et al. on 316 healthcare workers who had contact to COVID-19 patients in a tertiary hospital in Essen, Germany found a SARS-CoV-2-IgG antibody seroprevalence of 1.6% between March 25th and April 21th, 2020.^31^ This may be partly explained by the different epidemiological situation in the respective countries. Austria had a low incidence during sample collection of this study with a 14-day notification rate of reported cases per 100 000 population ranging between 0.0 and 88.2 during the study period (May 11^th^ – August 23^rd^).^32^ Additionally, the occupational risk was reduced in the participating trauma hospitals/rehabilitation centers because infected patients have been transferred to specifically implemented corona wards in other hospitals in Austria.

### Asymptomatic cases

Among participants who had positive test results in either of the antibody tests, 50.4% did not report any symptoms consistent with common manifestations of COVID-19 (fever, fatigue, dry cough, dyspnea, nasal congestion, headaches, myalgia, or diarrhea ^1,9,33–35^) in the preceding weeks irrespective of their age. Previous literature reported that around 30% of seropositive participants were asymptomatic.^21,30,36^ Antibody levels were lower in smokers and in women who had a less severe course of disease of COVID-19, and were generally elevated in older adults.^21^

### Sensitivity and specificity of antibody tests

Sensitivity and specificity rates of different SARS-CoV-2 antibody tests vary between 53% and 94%, and between 91% and 99.5%, respectively.^37^ Due to the pandemic, an urgent need for outpatient and rapid field detection of antibodies against SARS-CoV-2 virus emerged. Therefore, a valid antibody test it is of utmost interest for society and previous literature has reported about validity of several rapid antibody tests:

Li et al. used the SARS-CoV-2 rapid IgG-IgM combined antibody test kit to detect IgM and IgG antibodies against SARS-CoV-2 virus in human blood. The study reported a sensitivity of 88.66%, and a specificity of 90.63%, considering a positive test result if IgM, or IgG, or both are positive. In their study, 64.48% (256 out of 397) of positive patients had both IgM and IgG antibodies 8-33 days after infection symptoms appeared, and test results from venous blood and fingertip blood matched with 100% consistency.^26^ Another study by Lee and colleagues used the ALLTEST 2019-nCoV IgG/ IgM Rapid Test Cassette and found a sensitivity of 100%, a specificity of 98.0%, and an accuracy rate of 98.6% for the anti-SARS-CoV-2 IgG antibody. IgG appeared at post-exposure 18-21 days or the illness day 11. The relative sensitivity for the IgM antibody was reported to be 85% (95% CI, 62.1% - 96.8%) ^27^ In contrast, sensitivity rates of the antibody lateral flow assay in our study showed a wide variation ranging between 36.0% and 69.2% depending on the type of sampling (whole blood or serum) and the time point of sample collection (T1 vs. T2) in the group who had tested positive via PCR prior to study inclusion. Surprisingly, sensitivity rates were higher in participants with a positive antibody test during the study and no positive PCR prior to the study, ranging between 83.0% and 100%. The rapid antibody test generally shows higher sensitivity when based on serum than on whole blood.

Summarizing these results, the rapid antibody test based on whole blood can - at best - be used for a pre-screening for suspected cases. Based on serum, the test showed better sensitivity which may be due to higher antibody concentration in serum versus whole blood..

In our study, the ELISA tests showed an overall sensitivity of 88.9% in the Roche test and 81.5% in the Diasorin test using the neutralization test as a reference.

### Antibody production and waning

Guo et al. conducted a longitudinal study on 82 confirmed and 58 probable cases using a nucleocapsid-based ELISA and found that the production of IgM, IgA and IgG antibodies against SARS-CoV-2 were positive as early as day one after symptom onset. The median duration of IgM and IgA antibody detection were 5 days (IQR 3-6), while IgG was detected 14 days (IQR 10-18) after symptom onset. IgM antibody levels increased between days 8– 14; but did not increase further between days 15–21, or after day 21. The IgG antibodies could be detected on days 0–7, and continued to increase on days 8–14, as well as days 15-21, and plateaued by day 21.^16^

Previous literature reported, that antibodies remained stable for four months after diagnosis ^21,36^ In our study, a significant decline in neutralizing antibody concentration was observed between time points of testing within a period of approximately six months, but PRNT tests remained well above threshold in most patients indicating persistence of neutralizing antibodies. This result supports national vaccination recommendations in post-COVID patients which recommend suspending vaccination in such individuals.^38,39^ This is in contrast to US recommendations (no suspension of vaccination).^40^

### Limitations

Antibody seroprevalence may be underestimated if participants had not yet produced a sufficient antibody response at the time of sample collection, or if antibody titers had already declined since the infection. We tried to reduce this bias by conducting the PCR and LFA tests twice with approximately six weeks in between testing (between T1 and T2) and an additional serum sample collection for ELISA / PRNT after another three months (between T2 and T3).

## Conclusion

This study identified the following factors related to SARS-CoV-2 infection among health care workers in Austrian trauma centers and rehabilitation facilities: During the study period (May 11^th^ – December 21^th^) only 0.81% were tested positive for neutralizing antibodies, indicating that non-pharmaceutical interventions and other epidemiological interventions as recommended in Austria at the time of the study were widely effective in keeping infection rates low.

The antibody concentration in patients tested positive for COVID decreases significantly over a period of six months with 14.8% (4 out of 27) losing detectable antibodies.

Also, the results of rapid antibody tests are questionable due to a wide variability in their sensitivity rates.

## Data Availability

Data can be made available upon reasonable request to the corresponding author.

## Funding

The project was funded by the Austrian Social Insurance for Occupational Risks (AUVA) and the authors would like to express their sincere appreciation.

## Ethical Considerations and consent to participate

All procedures performed in this study were in accordance with the ethical standards of the institutional and national research committee and with the 1964 Helsinki declaration and its later amendments or comparable ethical standards. All participants provided written, informed consent. The study was approved by the ethics committee of the Austrian social insurance for occupational risks (AUVA) (Vote No. 04/2020). The study was registered on the ClinicalTrials.gov database (unique identifier: **NCT04354779)**.

All authors have read and approved the manuscript.

## Notes

### Competing Interest Statement

The authors have declared no competing interest.

### Clinical Trial

NCT04354779

### Author Declarations

The study was approved by the ethics committee of the Austrian social insurance for occupational risks (AUVA) (Vote No. 04/2020).

